# Estimating enteric fever seroincidence in Bangladesh using rapid serosurveys

**DOI:** 10.64898/2026.01.11.26343883

**Authors:** Sira Jam Munira, Naito Kanon, Nahidul Islam, Md. Shakiul Kabir, Anik Sarkar, Shamsul Alam Polash, Hafizur Rahman, Alice S. Carter, Richelle C. Charles, Jason R. Andrews, Stephen P. Luby, Kristen Aiemjoy, Denise O. Garrett, Samir K Saha, Jessica C. Seidman, Senjuti Saha

**Author notes:** These authors contributed equally to this article (Sira Jam Munira, Naito Kanon). These authors contributed equally to this article (Denise O. Garrett, Samir K Saha, Jessica C. Seidman, Senjuti Saha). **Address for correspondence**: Senjuti Saha; Child Health Research Foundation, SEL Huq Skypark, 23/2 Khilji Road, Block-B, Mohammadpur, Dhaka-1207, Bangladesh;, Jessica C. Seidman; Sabin Vaccine Institute, 2000 Pennsylvania Ave, NW, Suite 7000, Washington, DC 20006, US.

## Abstract

**Background:** Enteric fever, caused by *Salmonella* Typhi or Paratyphi, is a major public health issue in low-and middle-income countries. Accurate burden estimation is hampered by limited microbiological facilities and low sensitivity of blood culture tests. Serosurveillance offers a scalable alternative to address these challenges. This study estimated the seroincidence of enteric fever in Bangladesh using cross-sectional rapid serosurveys.

**Methods:** School-based surveys (January-June 2022) were conducted in Chattogram, Dinajpur, Sylhet, Satkhira, and Faridpur, and community-based surveys (July 2019-November 2021) were conducted in Dhaka and Mirzapur. Blood samples (dried blood spots or venous blood) were collected and tested for anti-Hemolysin E IgG responses using a kinetic Enzyme-linked Immunosorbent Assay. Seroincidence was estimated using a maximum likelihood approach based on peak antibody titers and decay rates, modelled from blood-culture confirmed enteric fever cases in Bangladesh.

**Findings:** A total of 2,969 participants aged 0-22 years were enrolled, with 75.5% aged 5-15 years. Seroincidence rates (per 100 person-years) were highest in Dhaka (33.1; 95% confidence interval [CI], 29.0-37.9), followed by Mirzapur (19.5; 95%CI, 17.4-21.7) and Chattogram (17.1; 95% CI, 15.0-19.4), and the lowest in Faridpur (11.1; 95%CI, 9.6-12.7). The under 5 age group exhibited higher rates in Dhaka (40.3; 95%CI, 32.1-50.7) and Mirzapur (26.1; 95%CI, 21.0-32.4) compared to older age groups. Seroincidence was elevated in areas with higher population density.

**Conclusions:** This study highlights a substantial burden of enteric fever infections across Bangladesh. These provide important evidence to guide post-introduction monitoring and optimization of the typhoid conjugate vaccine program in the national immunization schedule.

**Key points:** This study provides the first multi-site, rapid seroincidence estimates of typhoid and paratyphoid fever across diverse urban and rural settings in Bangladesh, revealing a substantial and previously under-recognized burden, particularly among young children. By measuring antibody responses using a targeted serologic blood test and applying a novel analytical approach based on Bangladesh-specific antibody kinetics to cross-sectional school- and community-based surveys, it addresses critical gaps left by blood culture–based surveillance, which is limited in coverage and sensitivity. Our findings demonstrate that scalable serosurveillance can reliably capture population-level transmission intensity where routine diagnostics are unavailable. These results directly inform post-introduction monitoring and optimization of the typhoid conjugate vaccine program and provide a practical framework for burden estimation and vaccine policy decision-making in other low-resource settings.

## INTRODUCTION

Enteric fever, caused by *Salmonella* enterica serovar Typhi and Paratyphi A, B, or C, remains a significant cause of mortality and morbidity in low- and middle-income countries [1]. Globally, it accounts for an estimated 9.2 million cases and 110,000 deaths annually, with South Asia bearing the highest burden (379/100,000 person-years) [2]. Dhaka, the capital of Bangladesh, has one of the highest reported clinical incidence rates globally, with an estimated 913/100,000 person-years for typhoid and 128/100,000 person-years for paratyphoid [3]. Clinical surveillance data show the highest incidence among children under 5 years, particularly those aged 2-4 years, followed closely by school-aged children [3,4].

Prior studies of typhoid incidence in Bangladesh have reported blood culture-based incidence data primarily from Dhaka, where microbiological laboratory facilities for blood culture, the reference standard for diagnosis, are available at multiple institutions [3,5]. Blood culture has limited sensitivity (∼60%) due to factors such as low pathogen load and prior antibiotic use [6]. Accurately estimating population-level incidence requires continuous surveillance and broad access to health facilities equipped for blood culture, both of which still remain key challenges. Without comprehensive surveillance, incidence estimates across the country may be incomplete, hampering evidence-based policy decisions like vaccine prioritization [7].

Serosurveillance has emerged as a practical and scalable tool to estimate population-level infection burden, particularly in resource-constrained settings. This approach has been successfully used for infectious diseases such as COVID-19, dengue, and hepatitis E [8–11]. Historically, the Vi capsular polysaccharide antigen was used as a serologic marker for typhoid fever. However, Vi has several drawbacks: it cannot distinguish between natural infection and Vi-based vaccination, performs poorly in detecting acute infections and paratyphoid cases, and does not show age-related variation in endemic settings [12–14]. Recent studies have identified *Salmonella* Typhi Hemolysin E (HlyE), a pore-forming toxin, as a promising serological marker for exposure to enteric fever [15,16]. These markers have also been further validated for use in population-based serosurveillance for enteric fever, demonstrating a strong correlation with clinical incidence estimates [17]. Previous studies utilizing these serological markers and employing a representative population-level sampling approach have reported seroincidence estimates in Bangladesh, Nepal, Pakistan, Ghana, South Sudan, India, and Kenya [17–20].

To address gaps in clinical surveillance and better understand the burden of enteric fever across diverse regions of Bangladesh, this study aimed to estimate seroincidence using a school-based sampling approach. This sampling strategy could be a practical approach to conduct serosurveys rapidly and efficiently. We also incorporated a mix of urban, semi-urban, and rural regions to evaluate the diversity of enteric fever burden and the pattern of infection nationally. These findings are intended to inform evidence-based policies for the strategic implementation and post-introduction monitoring of the typhoid conjugate vaccine in Bangladesh.

## METHODS

### Study design, sites, and population

This cross-sectional study estimated the seroincidence of enteric fever across seven geographically diverse regions in Bangladesh. We conducted school- and community-based serosurveys to capture representative data from urban, semi-urban, and rural populations, including participants aged 0-22 years. For community-based surveys in Dhaka and Mirzapur, we randomly selected sampling grid clusters within specific catchment areas, where prior healthcare utilization surveys and ongoing demographic surveillance had been conducted [21,22]. All households in each selected cluster were enumerated, and an age-stratified random sample of participants aged 0-18 years was drawn. Details of catchment areas and randomization procedures for selecting sampling grids, households, and participants have been published previously [3,17,21]. Inclusion criteria were age (0-18 years) and residence within catchment areas. Participants were enrolled between July 2019 and November 2021.

For school-based surveys, we selected eight schools from four districts — Chattogram, Dinajpur, Sylhet, and Satkhira — and enrolled participants aged 4-18 years. Additionally, we included four project schools in Faridpur and enrolled participants aged 1-22 years old. These project schools served local communities by offering day care services, meals, and education. All surveys were conducted between January and June 2022. We selected districts representing urban, rural, and semi-urban communities across the central, northeastern, northwestern, southwestern, and southeastern regions of Bangladesh. Population densities were estimated using GPS location data collected during enrolment [23]. The schools were selected based on ease of transportation, logistical feasibility, and existing collaborations with the school authorities and local governing bodies. Within schools, participants were randomly selected from student registries. Teachers notified parents/guardians in advance about their child’s selection and requested informed consent. A study team comprising 13-15 members completed each survey within 3-4 days (Figure 1 and Supplementary Table 1).

**Figure 1.**
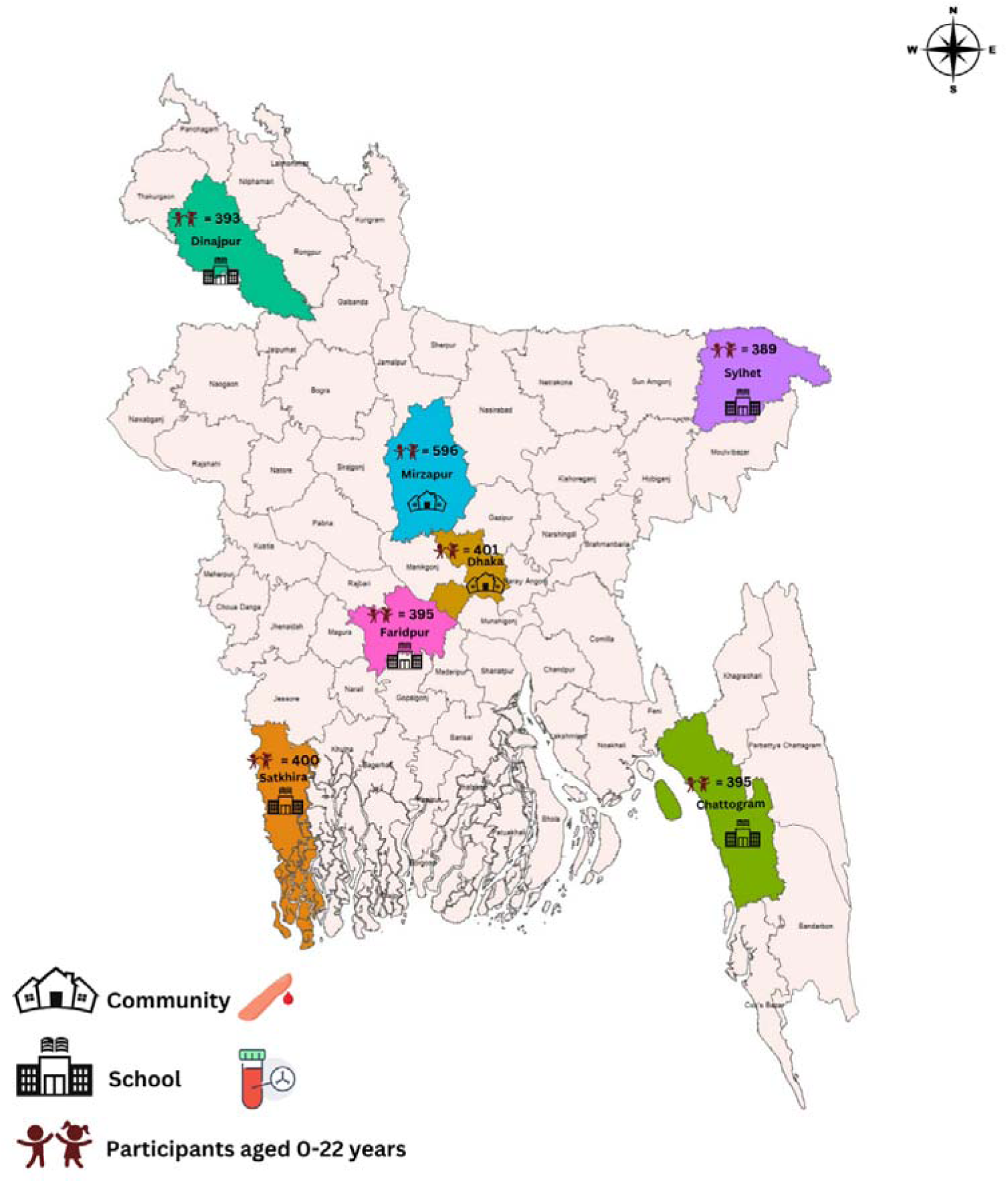
Overview of study areas, sampling procedure, and characteristics of study participants.

### Data collection

The study team administered a pre-tested, semi-structured questionnaire to collect socio-demographics, including age and sex, as well as health-related information such as self-reported episodes of fever in the past six months, typhoid diagnoses within the last year, and any history of typhoid vaccination. In community-based surveys, trained health workers administered questionnaires to parents/guardians during pre-scheduled home visits. In school-based surveys, the study team collected sociodemographic information from students during sample collection, when applicable. Later, they verified through follow-up phone calls with parents/guardians to ensure completeness and accuracy.

### Sample collection, storage, and laboratory procedure

Blood samples were collected using two different methods, based on the survey settings and logistical considerations (Figure 1). In the community-based serosurveys, finger stick capillary blood samples were collected onto TropBioTM filter paper (Cellabs, Brookvale, NSW, Australia) and stored as dried blood spots (DBS). Samples were air-dried for at least 2 hours at room temperature, sealed in air-tight plastic bags with desiccant, and transported to the central laboratory, where they were stored at -20°C until further processing. In school-based surveys, venous blood samples were collected at the request of the schools to include screening tests (blood grouping and Hepatitis B/C testing) to benefit the participants and local communities. Using aseptic technique, 2-3 mL of venous blood was collected in EDTA (Ethylenediamine tetraacetic acid) tubes. Approximately two-thirds of each blood sample was centrifuged to obtain plasma, while the remaining one-third was preserved in EDTA as whole blood. All samples were initially stored at 4°C locally and transported to our central laboratory in Dhaka within 72 hours, ensuring the temperature was maintained throughout. Whole blood was used for blood grouping, and plasma for Hepatitis B or C testing. After completion of the tests, the plasma samples were preserved at -20°C until further processing.

IgG antibody responses against HlyE were measured using kinetic enzyme-linked immunosorbent assays (K-ELISAs) and normalized to positive controls. Descriptions of DBS elution protocol, ELISA protocol, and serological analysis are available in previous publications [16,17,24] and in Supplementary Document.

### Statistical analysis

Our target sample size was at least 400 per area, as detailed in the supplementary materials. Seroincidence rates were estimated based on quantified anti-HlyE IgG antibody responses using a likelihood function constructed to model the observed cross-sectional antibody data. Incident infections were assumed to occur as a Poisson process. The likelihood function represents each individual’s antibody measurement as the product of antibody responses following infection, measurement variability, and baseline biological reactivity. Antibody responses after infection, including peak responses and decay rates, were parameterized using a previously published longitudinal cohort of blood-culture-confirmed enteric fever cases enrolled in Nepal, Pakistan, and Bangladesh [17], and incorporated directly into the likelihood to characterize expected temporal antibody trajectories following infection. Measurement error was quantified using the coefficient of variation across assay replicates, and biological noise was defined using responses among confirmed unexposed negative controls [17,18,25]. For this study, we used unstratified and age-stratified antibody decay dynamics modelled from a subset of 407 blood-culture confirmed cases enrolled in Bangladesh [26]. Seroincidence estimates were calculated for each of the seven study sites. Age-stratified analyses were conducted for three groups: under 5 years, 5-15 years, and 16 years and older. We also conducted a sensitivity analysis using pooled multi-country kinetics to assess differences in seroincidence estimates [17].

All statistical analyses were performed using R version 4.2.2 and the serocalculator package 1.3.0 [27]. Confidence intervals for seroincidence estimates were computed at the 95% level.

## RESULTS

### Study population characteristics

A total of 2,969 participants were enrolled across seven regions of Bangladesh (Figure 1). 997 (33.6%) were from community-based surveys, and 1972 (66.4%) were enrolled in school-based surveys. The majority of the participants (2243/2969, 75.5%) were aged 5-15 years. The median age of participants varied by region, ranging from 9-14 years, and females constituted 54.9% (1630/2969) of the participants. About 12.5% (371/2969) of participants reported having a fever within the past six months, and only 2.8% (84/2969) reported a prior diagnosis of typhoid fever. Most participants (2937/2969, 98.9%) reported never receiving a typhoid vaccine in their lifetime (Table 1).

**Table 1.** Characteristics of the study participants.

| Characteristics | Dhaka<br>(n=401) | Mirzapur<br>(n=596) | Chattogram<br>(n=395) | Dinajpur<br>(n=393) | Sylhet<br>(n=389) | Satkhira<br>(n=400) | Faridpur<br>(n=395) |
| --- | --- | --- | --- | --- | --- | --- | --- |
| **Median age in years (IQR)** | 9.2 (4.9-14) | 9.9 (4.9- 14.5) | 10 (9-13) | 11 (9-13) | 14 (12-16) | 13 (11-15) | 9 (6-14) |
| **Age categories** |  |  |  |  |  |  |  |
| **Under 5 years (%)** | 101 (25.2%) | 151 (25.3%) | 0 (0.0%) | 2 (0.5%) | 1 (0.3%) | 0 (0.0%) | 62 (15.7%) |
| **5-15 years (%)** | 256 (63.8%) | 360 (60.4%) | 380 (96.2%) | 372 (94.7%) | 283 (72.8%) | 347 (86.8%) | 245 (62.0%) |
| **16 years and older (%)** | 44 (11.0%) | 85 (14.3%) | 15 (3.8%) | 19 (4.8%) | 105 (26.9%) | 53 (13.3%) | 88 (22.3%) |
| **Female (%)** | 218 (54.4%) | 299 (50.2%) | 282 (71.4%) | 213 (54.2%) | 186 (47.8%) | 241 (60.3%) | 191 (48.4%) |
| **Self- reported fever episodes in the past six months** |  |  |  |  |  |  |  |
| **Yes (%)** | 40 (10.0%) | 121 (20.3%) | 43 (10.9%) | 40 (10.2%) | 42 (10.8%) | 45 (11.3%) | 40 (10.1%) |
| **No (%)** | 361 (90.0%) | 475 (79.7%) | 350 (88.6%) | 252 (64.1%) | 346 (88.9%) | 354 (88.5%) | 322 (81.5%) |
| **Don't know (%)** | 0 (0.0%) | 0 (0.0%) | 2 (0.5%) | 101 (25.7%) | 1 (0.26%) | 1 (0.3%) | 33 (8.4%) |
| **History of typhoid diagnosis** |  |  |  |  |  |  |  |
| **Yes (%)** | 25 (6.2%) | 4 (0.7%) | 16 (4.1%) | 14 (3.6%) | 7 (1.8%) | 6 (1.5%) | 12 (3.0%) |
| **No (%)** | 376 (93.8%) | 592 (99.3%) | 378 (95.7%) | 283 (72.0%) | 381 (97.9%) | 389 (97.3%) | 355 (89.9%) |
| **Don't know (%)** | 0 (0.0%) | 0 (0.0%) | 1 (0.3%) | 96 (24.4%) | 1 (0.3%) | 5 (1.3%) | 28 (7.1%) |
| **No typhoid vaccination (%)** | 371 (92.5%) | 596 (100%) | 395 (100%) | 393 (100%) | 388 (99.8%) | 399 (99.8%) | 395 (100%) |

\*IQR, Interquartile range.

### Regional seroincidence

Anti-HlyE IgG responses demonstrated substantial regional and age-related variation (Figure 2). Overall antibody response was highest in Dhaka with a median of 9.3 ELISA units and an interquartile range (IQR) of 4.9-18.9, followed by Chattogram (6.3; IQR, 3.4-10.2) and Mirzapur (5.4; IQR, 3.1-9.1), while they were comparatively lower in Faridpur (3.7; IQR, 1.8-7) (Figure 2A). These differences in responses were statistically significant (p<0.0001) across all areas compared to Dhaka. Antibody responses increased with age in all seven regions. Dhaka consistently exhibited the highest responses amongst all ages (Figure 2B).

**Figure 2.**
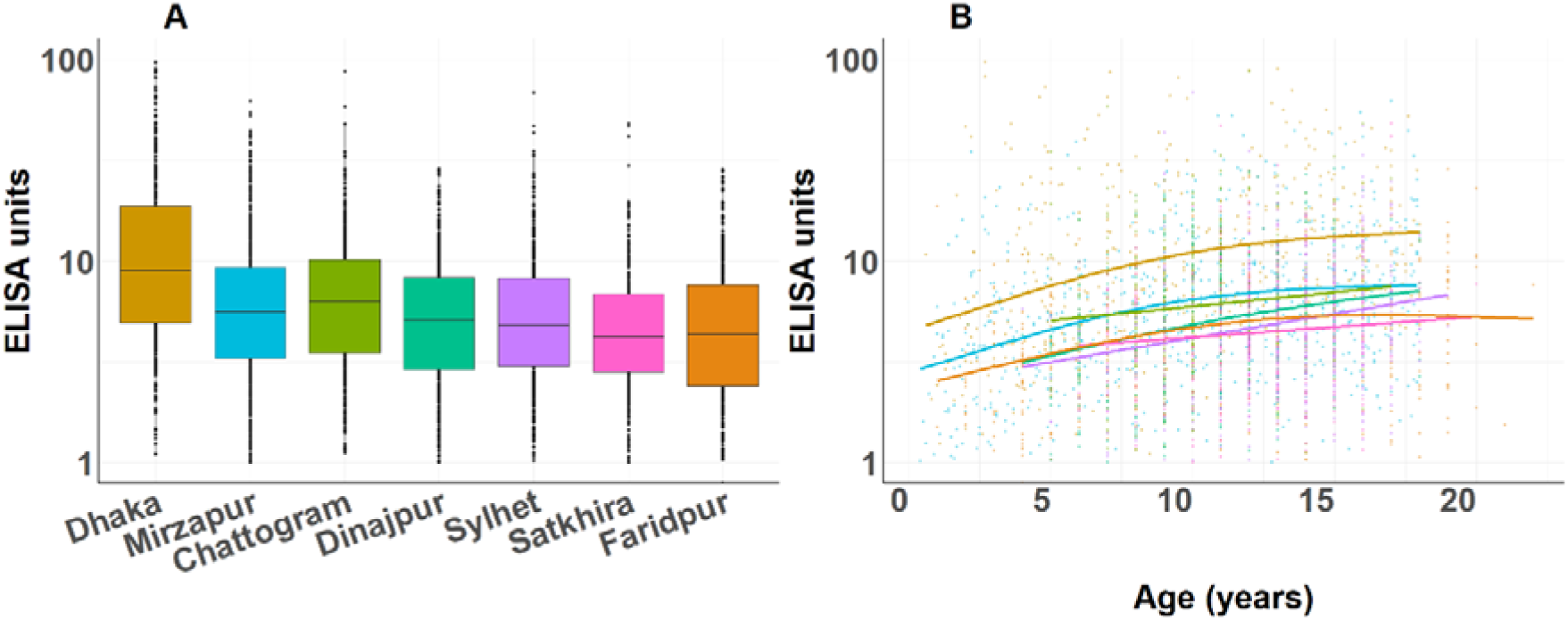
Anti-HlyE IgG responses among study participants A) by study areas B) by age. Each dot represents each participant’s anti-Hemolysin E IgG responses results; Box plots represent median responses with the Interquartile range (IQR) across areas.

The overall seroincidence (per 100 person-years) was highest in Dhaka (33.1; 95%CI, 29.0-37.9) and then in Mirzapur (19.5; 95%CI, 17.4-21.7), both of which were assessed through community-based surveys. In comparison, school-based surveys conducted in the remaining areas reported lower seroincidence estimates: Chattogram (17.1; 95%CI, 15.0-19.4), Dinajpur (14.4; 95%CI, 12.6-16.3), Sylhet (12.6; 95%CI, 11.0-14.4), Satkhira (11.5; 95%CI, 10.1-13.0), and Faridpur (11.1; 95%CI, 9.6-12.7) (Table 2, Figure 3). While differences in survey approach may have contributed, these patterns also align with population density, with higher seroincidence observed in more densely populated areas (r=0.87) (Supplementary Figure 1).

**Figure 3.**
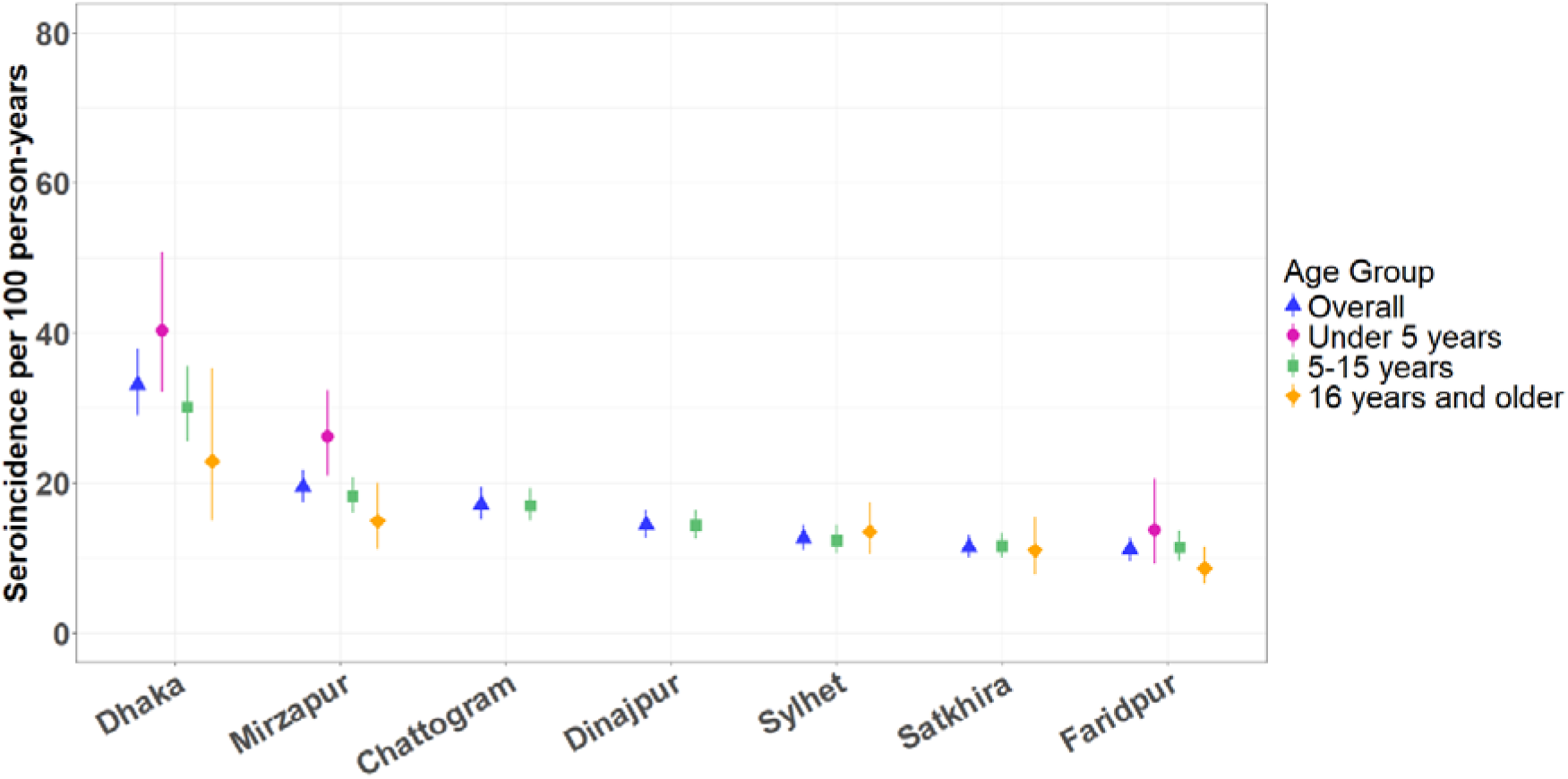
Estimated seroincidence of enteric fever per 100 person-years by age groups across seven areas. Each shape represents a seroincidence estimate with a 95% confidence interval, presented both overall and stratified by age group.

**Table 2.** Estimated seroincidence of enteric fever in Bangladesh.

| Study areas | Seroincidence per 100 person-years* (95% CI**) |  |  |  |
| --- | --- | --- | --- | --- |
|  | Overall | Under 5 years | 5-15 years | 16 and older years |
| **Dhaka** | 33.1 (29.0- 37.9) | 40.3 (32.1- 50.7) | 30.1 (25.5- 35.5) | 22.9 (14.9- 35.3) |
| **Mirzapur** | 19.5 (17.4- 21.7) | 26.1 (21.0- 32.4) | 18.2 (16.0- 20.8) | 14.9 (11.1- 20.0) |
| **Chattogram** | 17.1 (15.0- 19.4) | Not applicable | 17.0 (14.9- 19.3) | Not applicable |
| **Dinajpur** | 14.4 (12.6- 16.3) | Not applicable | 14.3 (12.5- 16.3) | Not applicable |
| **Sylhet** | 12.6 (11.0- 14.4) | Not applicable | 12.3 (10.6- 14.4) | 13.5 (10.5- 17.4) |
| **Satkhira** | 11.5 (10.1- 13.0) | Not applicable | 11.6 (10.1- 13.3) | 11.0 (7.8- 15.5) |
| **Faridpur** | 11.1 (9.6- 12.7) | 13.7 (9.1- 20.5) | 11.4 (9.7- 13.6) | 8.6 (6.5- 11.5) |

\*Bangladesh-specific pooled kinetics were used for the seroincidence estimation. Overall seroincidence estimates were generated using Bangladesh-specific unstratified antibody kinetics. Bangladesh-specific age-stratified kinetics were applied to generate estimates in all age groups.
\*\*95% CI, 95% confidence interval

### Stratified seroincidence by age, gender, and prior typhoid diagnosis

Seroincidence estimates per 100 person-years were available for children under 5 years in three areas. The highest burden was in Dhaka (40.3; 95%CI, 32.1-50.7), compared to Mirzapur (26.1; 95%CI, 21.0-32.4) and Faridpur (13.7; 95%CI, 9.1-20.5). For the 5-15 years age group, community-based sampling showed the highest burden in Dhaka (30.1; 95%CI, 25.5-35.5), followed by Mirzapur (18.2; 95%CI, 16.0-20.8). In the remaining areas, where a school-based sampling strategy was used, Chattogram had the highest seroincidence (17.0; 95%CI, 14.9-19.3), and Faridpur had the lowest (11.4; 95%CI, 9.7-13.6). For those 16 years and older, Dhaka (22.9; 95%CI, 14.9-35.3) and Mirzapur (14.9; 95%CI, 11.1-20.0) had the highest burden, while Faridpur had the lowest (8.6; 95%CI, 6.5-11.5). Seroincidence could not be estimated in this age group for Chattogram and Dinajpur due to insufficient sample size. Children under 5 consistently exhibited higher seroincidence rates than older age groups in high-burden settings. In contrast, lower-incidence areas had more uniform age distributions, suggesting potential differences in exposure patterns or transmission intensities (Table 2, Figure 3).

Seroincidence was estimated using pooled multi-country kinetics for comparison with Bangladesh-specific kinetics. Estimates based on the Bangladesh kinetics are lower in all sites and age groups because they have a slower decay rate than those based on the pooled kinetics. In a high force of infection setting such as Dhaka, pooled kinetics may overestimate seroincidence (Supplementary Table 2 and Supplementary Figure 2).

Stratified seroincidence by gender was estimated across all sites combined and separately. Overall, seroincidence estimations were slightly higher among females (16.5; 95%CI, 15.5-17.6) compared to males (14.9; 95%CI, 13.9-16.1) in all areas. Gender-specific seroincidence estimates were highest in Dhaka (Female: 33.9; 95%CI, 28.2-40.8 and Male: 32.3; 95%CI, 26.5-39.3) and lowest in Faridpur (Female: 10.4; 95%CI, 8.5-12.7 and Male: 11.8; 95%CI, 9.7-14.3). Participants with a typhoid diagnosis in the last year showed higher seroincidence (20.5; 95%CI, 15.2-27.8) compared to those without a reported typhoid diagnosis (15.9; 95%CI, 15.1-16.7) (Supplementary Table 3).

## DISCUSSION

This study of 2,969 participants from seven geographically diverse regions of Bangladesh revealed a high enteric fever burden in areas where traditional blood culture surveillance data are not available. We collected plasma and fingerstick capillary blood samples through cross-sectional rapid school- and community-based surveys. While a population-based random sampling strategy is the gold standard for estimating disease burden, the school-based approach enabled low-cost, efficient sample collection with minimal logistical challenges. Additionally, the centralized nature of schools facilitated coordination with the study team during the initial preparation phase. Also, it ensured the smooth delivery of blood group and hepatitis test results to participants. This study identified substantial evidence of high enteric fever infections in rural and semiurban Bangladesh, particularly among children under 5 years, underscoring their vulnerability and the need for targeted interventions.

Our study estimated the incidence of enteric fever infections based on quantitative HlyE IgG antibody responses [17]. Our results show that while anti-HlyE IgG responses increased with age across all seven areas, the highest seroincidence rates were observed in the youngest age groups (children <5 years old). This finding is consistent with the Strategic Typhoid Alliance Across Asia and Africa (STRATAA) study in Bangladesh [28]. For the under 5 age group, our seroincidence estimates in Dhaka (40.3), Mirzapur (26.1), and Faridpur (13.7) are significantly higher compared to the older age groups. This finding suggests a high incidence of enteric fever among younger children, consistent with clinical incidence data [3,29]. The higher ratio of seroincidence to clinical incidence among young children observed here and in other seroepidemiology studies suggests that children may be more likely to have subclinical infections or that culture-confirmed infections are under-ascertained in this age group (due to the lower sensitivity of blood culture at lower volumes, for example).

Our seroincidence estimates were highly correlated with increased population densities. Dhaka, the most densely populated area, exhibited the highest seroincidence estimates per 100 person-years (33.1) among all seven regions of Bangladesh. However, previous seroincidence estimates from the SEES study (41.2), using anti-HlyE IgA and IgG responses, and STRATAA estimates (47.1) using HlyE IgG were higher than our finding [17,28].

Differences between our seroincidence estimate and that of the SEES study may reflect our use of Bangladesh-specific antibody kinetics, in contrast to the multi-country kinetics applied in SEES. In Bangladesh, IgG responses decayed more slowly in comparison to the other sites, likely explained by the higher force of infection there. When using Bangladesh-specific kinetics, time-since-infection at the population-level is longer and the seroincidence rate is therefore smaller. IgA responses are less dependent on different forces of infection and may be preferable to use in future studies to allow for more robust comparisons across sites.

Mirzapur had the second-highest seroincidence estimate (19.5), while a previous study based on clinical incidence, from 2016 to 2017, had reported it as a low-endemic area [30]. Changes over the years in population migration, improved transportation, and increasing urbanization may have influenced this shift by facilitating greater transmission of *Salmonella* Typhi/Paratyphi than reported historically. Alternatively, *Salmonella* Typhi/Paratyphi may be under detected in this area. Chattogram, the second largest city in Bangladesh, reflected the third-highest seroincidence estimate (17.1). A prior study at Chattogram found that one in five (52/300, 17.3%) non-malarial febrile patients admitted had a diagnosis of enteric fever, detected by blood culture or PCR [31]. All six rural and semi-urban areas in our study displayed considerably lower seroincidence estimates compared to Dhaka, an urban area. Although seroincidence estimates are expected to exceed clinical incidence, our findings across all seven regions and age groups still revealed high levels of enteric fever transmission, well above the clinical high burden threshold of 100 cases per 100,000 person-years [3,32]. The SEES study also showed the higher rank order of seroincidence estimates tracked with clinical enteric fever incidence estimates derived from a blood culture-based surveillance approach [17].

An important advantage of using the school-based approach is its logistical efficiency. While not fully representative of the general population, it may serve as a reasonable proxy for estimating incidence among school-aged children where clinical surveillance data are lacking. School-based sampling significantly reduces demands on time, personnel, and resources, eliminates the need for household enumeration, and reduces travel and operational costs. For example, we were able to sample a school within 3-5 days, rather than several weeks/months to conduct a population-based serosurvey. These efficiencies make school-based surveys a valuable and feasible alternative in resource-constrained environments.

This study is subject to several limitations. First, the schools were selected conveniently, which may have introduced potential selection bias and reduced the generalizability of the findings. While this approach enabled efficient implementation in resource-limited settings, the selection process may not reflect the full geographic or socio-economic diversity of Bangladesh. Future studies using population-based sampling methods can validate the representativeness and expand upon these findings. Second, the school- and population-based serosurveys were conducted at different times of the year, which may limit direct comparability; however, because the incidence of enteric fever remains high year-round in Bangladesh, this is unlikely to introduce substantial bias. Third, enteric fever cases for the longitudinal cohort were enrolled from a pediatric hospital, and there were no cases older than 18 years. However, we have observed that the biggest differences in antibody decay dynamics are among young children (under 5 years) and older than 5 years. The unavailability of longitudinal antibody kinetics data beyond 15 years of age compelled us to use the Bangladesh-specific parameters from 5 years and older aged participants to assess the seroincidence in the 16 years and older age group. More extensive study of antibody kinetics data across different age groups and geographic areas would provide a clearer understanding of anti-HlyE immune responses after infection. Fourth, the analysis relied on Bangladesh-specific HlyE IgG dynamics, although IgA may be a better marker for comparing across contexts. Because the HlyE IgG responses were previously found to vary by study site, likely due to differences in the force of infection, this study used a longitudinal antibody kinetics modelled from enteric fever cases in Bangladesh. Alternatively, using a marker that exhibits consistent peaks and decays across settings with different forces of infection, such as HlyE IgA, may be a more feasible approach to scale enteric fever serosurveillance. Finally, we did not implement both community- and school-based approaches within the same area, which limited our ability to compare the two sampling strategies directly. For better comparison and understanding of their relative effectiveness, future studies could consider evaluating both approaches in the same population.

## CONCLUSIONS

Our study highlights a substantial burden of enteric fever infections in semi-urban and rural areas of Bangladesh. We demonstrated that a school-based sampling is a cost-effective method for rapidly determining the seroepidemiology of enteric fever. Seroincidence estimates from this method offer valuable insights into the national landscape of enteric fever burden. Since Bangladesh introduced the typhoid conjugate vaccine into the routine immunisation program on 12 October 2025, this approach can also help guide post-introduction monitoring of typhoid and paratyphoid trajectories among children in Bangladesh.

## NOTES

## Acknowledgements

We express our gratitude to the parents and teachers of all participants included in this study, as well as to the field and laboratory staff of the Child Health Research Foundation (CHRF) in Bangladesh. This study could not have been implemented without the support of local district authorities. We also acknowledge the collaborative efforts of the SIDRAH Foundation, Dhaka, Bangladesh, and Mr Kinkar Ghosh for the successful completion of this project.

## Author contributions

SS, SKS, JRA, SPL, and DOG conceptualized the study and funding acquisition. SJM, SS, and SKS formulated the methodology and implementation plan. All authors did the investigation. SJM, MSK, SAP, and SS supervised enrollment, data collection, and sample collection. NI, AS, and HR conducted and monitored all laboratory procedures. NI, JCS, and KA did all serological analyses. NK, SJM, ASC, KA, and JCS did data management, analysis, and visualization. SJM, SS, JCS, and NK drafted the manuscript. SJM, NK, SPL, RCC, JRA, KA, DOG, SKS, JCS, and SS critically analysed and revised the manuscript. NK, SJM, JCS, and KA have verified the data. All authors reviewed the manuscript and agreed with its contents.

## Patient consent statement

Informed consent was obtained from the parents or guardians of all participants. Participants aged 16 to <18 years provided their assent in addition to parental consent. For the community-based surveys, informed written consent was obtained during home visits. For the school-based surveys, school authorities obtained verbal consent from parents or guardians, and the study team communicated later with them by phone to obtain informed verbal consent. This study was approved by the ethical review board of the Bangladesh Shishu Hospital and Institute [(BICH-ERC-01/02/2019) and (BICH-ERC-09/02/2021)].

## Data availability statements

All data and analysis codes are available to the corresponding authors upon request.

## Supplementary Materials

This supplement provides additional details on sample size calculation, DBS elution protocol, Enzyme-linked Immunosorbent Assay (ELISA) protocol, measurement error, and biological noise estimation. It also includes information on study sites, seroincidence stratified by population density, gender, and prior typhoid history, and a comparison of seroincidence estimates using multi-country versus Bangladesh-specific kinetics.

## SUPPLEMENTARY DOCUMENT

### Sample size calculation

Our targeted sample size in Dhaka was 400 participants to estimate the seroincidence rate per 100 person-years in a 95% confidence interval (CI). Assuming an anticipated seroincidence of 25 cases per 100 person-years, this sample size would provide a precision of approximately ±4.9, corresponding to a relative precision of ±20% under a Poisson model. In Mirzapur, we estimated a larger sample size (∼600) because of an anticipated low seroincidence. In the remaining areas, 400 participants per district were enrolled to ensure sufficient power for estimating regional seroincidence rates. Further details were described elsewhere [1].

### ELISA methods

For DBS samples, two filled protrusions from the filter paper were cut and immersed in 133 μL of 1×PBS containing 0.05% Tween buffer. The samples were incubated overnight at 4°C, and eluates were collected following centrifugation. Plasma samples were processed directly without elution.

Microplates were coated with purified HlyE (1 µg/mL) as described in earlier studies [2,3]. Samples were added to the plates in duplicates, with plasma diluted at 1:5000 and DBS diluted at 1:500, accounting for the DBS eluate being 10 times more diluted than the plasma. Bound primary antibodies were detected using horseradish peroxidase (Jackson ImmunoResearch) conjugated goat anti-human IgG. Peroxidase activity was measured using the chromogenic substrate, ortho-phenylene diamine, and read kinetically at 450 nm for five minutes at 19-second intervals [4]. To minimize inter-plate and inter-site variability, the blank-adjusted sample readings were averaged, divided by the standard readings, which were included on each plate (Human chimeric monoclonal antibody), multiplied by 100, and expressed as ELISA units.

### Measurement error, biologic noise, and lower limit of detection for seroincidence estimation

We accounted for measurement error (assay variability) and biologic noise (background responses in unexposed individuals) as described in the SEES study [1]. Measurement error was quantified using the coefficient of variation (CV) from 100 positive and negative controls tested in triplicate at each site [5]. Biologic noise was estimated from stored serum samples of 48 children (1-5 years) with celiac-affected relatives and 31 healthy controls (2-18 years) from two celiac disease studies [6]. Lower limits of detection were determined by serial dilutions, identifying the point where CV ≥30%, adding two standard deviations, and normalizing to positive controls [7]. These thresholds were calculated separately for each laboratory.

---

**Supplementary Table 1.**
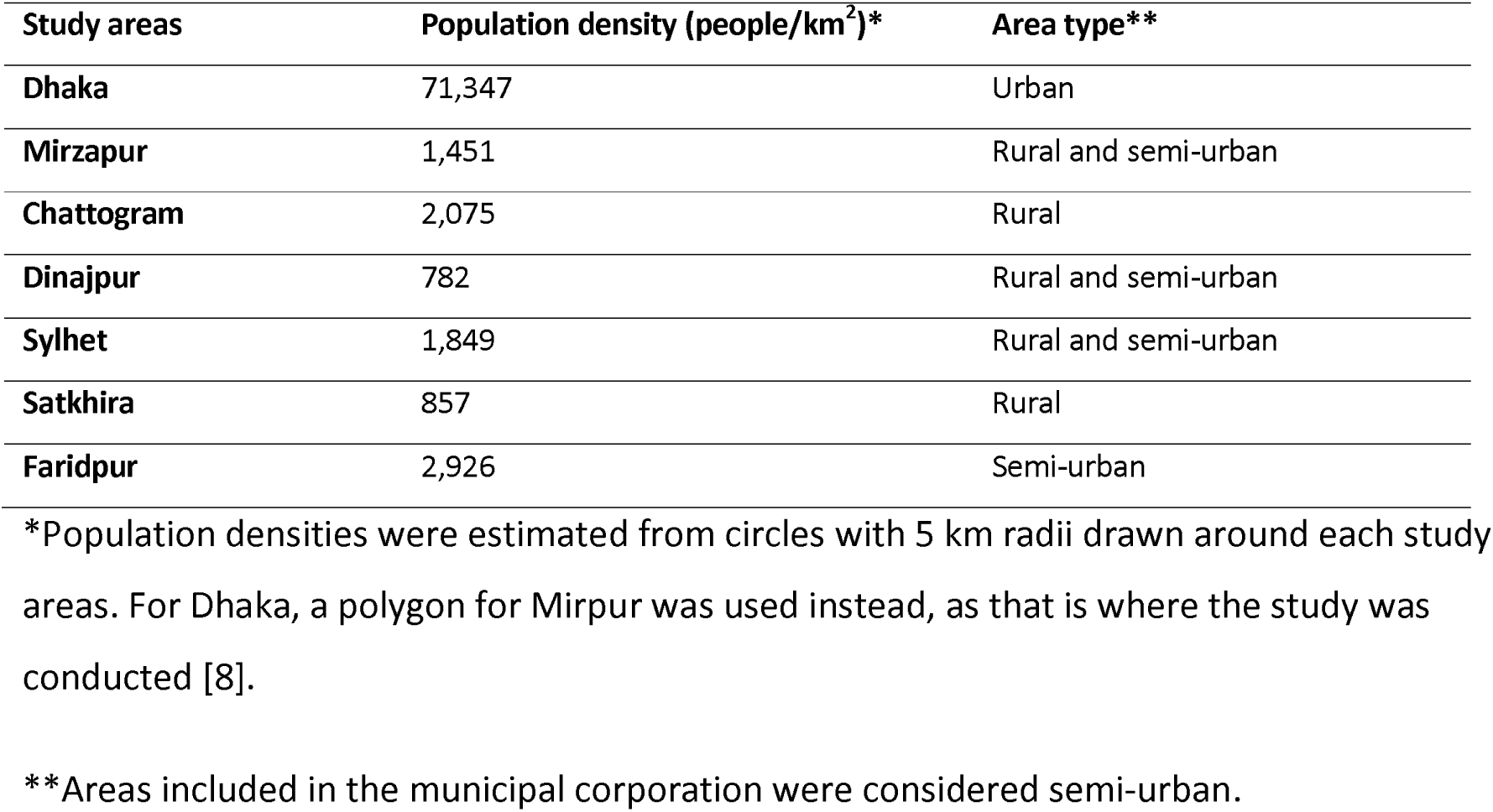
Study sites.

| Study areas | Population density (people/km<sup>2</sup>)* | Area type** |
| --- | --- | --- |
| Dhaka | 71,347 | Urban |
| Mirzapur | 1,451 | Rural and semi-urban |
| Chattogram | 2,075 | Rural |
| Dinajpur | 782 | Rural and semi-urban |
| Sylhet | 1,849 | Rural and semi-urban |
| Satkhira | 857 | Rural |
| Faridpur | 2,926 | Semi-urban |

\*Population densities were estimated from circles with 5 km radii drawn around each study areas. For Dhaka, a polygon for Mirpur was used instead, as that is where the study was conducted [8].
\*\*Areas included in the municipal corporation were considered semi-urban.

**Supplementary Table 2.**
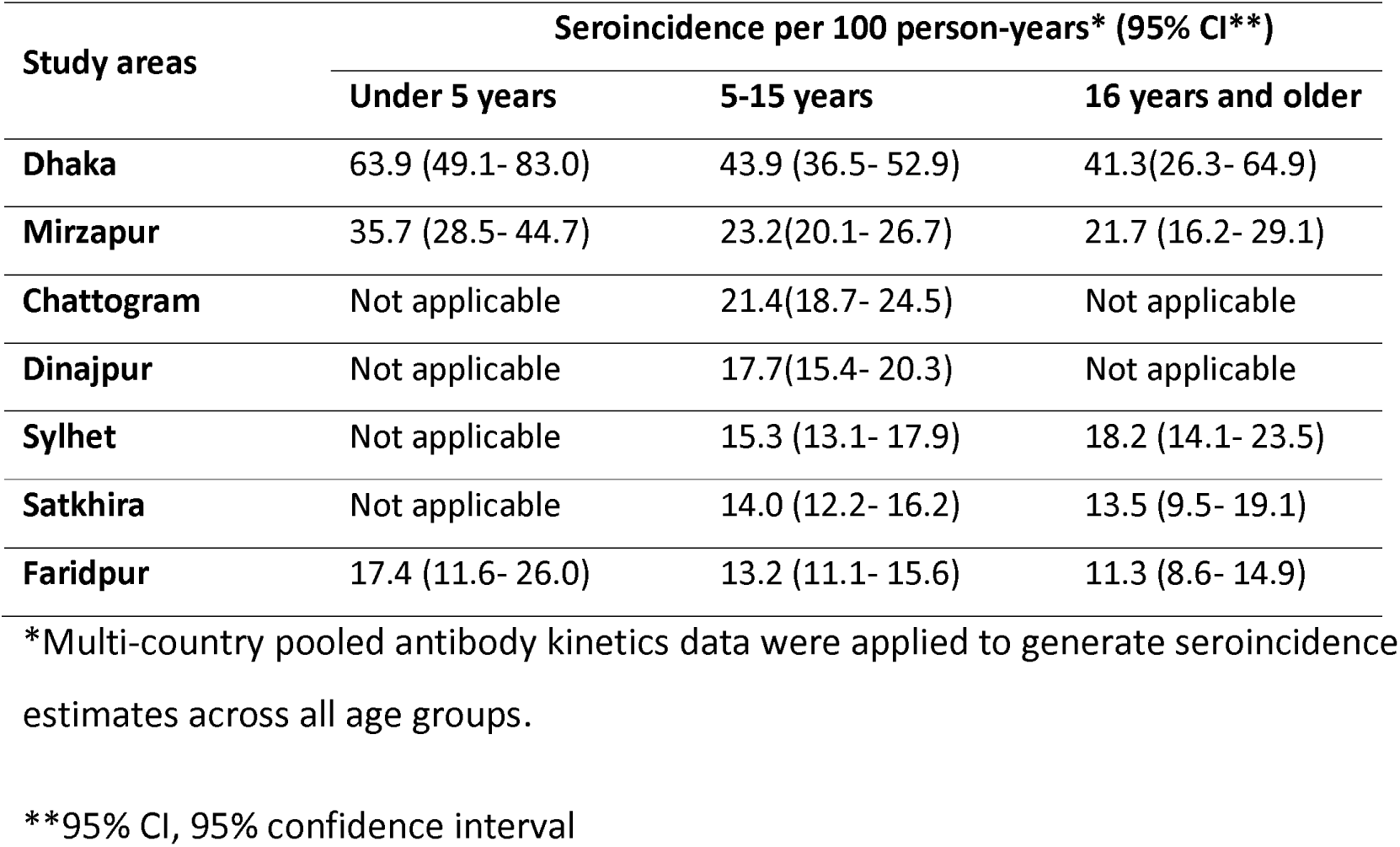
Estimated age-stratified seroincidence of enteric fever in Bangladesh using multi-country pooled kinetics.

| Study areas | Seroincidence per 100 person-years* (95% CI**) |  |  |
| --- | --- | --- | --- |
|  | Under 5 years | 5-15 years | 16 years and older |
| **Dhaka** | 63.9 (49.1- 83.0) | 43.9 (36.5- 52.9) | 41.3(26.3- 64.9) |
| **Mirzapur** | 35.7 (28.5- 44.7) | 23.2(20.1- 26.7) | 21.7 (16.2- 29.1) |
| **Chattogram** | Not applicable | 21.4(18.7- 24.5) | Not applicable |
| **Dinajpur** | Not applicable | 17.7(15.4- 20.3) | Not applicable |
| **Sylhet** | Not applicable | 15.3 (13.1- 17.9) | 18.2 (14.1- 23.5) |
| **Satkhira** | Not applicable | 14.0 (12.2- 16.2) | 13.5 (9.5- 19.1) |
| **Faridpur** | 17.4 (11.6- 26.0) | 13.2 (11.1- 15.6) | 11.3 (8.6- 14.9) |

\*Multi-country pooled antibody kinetics data were applied to generate seroincidence estimates across all age groups.
\*\*95% CI, 95% confidence interval

**Supplementary Table 3.** Estimated seroincidence of enteric fever in Bangladesh stratified by gender and history of typhoid diagnosis.

| Study Areas | Seroincidence per 100 person-years (95% CI**) |  |  |  |
| --- | --- | --- | --- | --- |
|  | Female | Male | Typhoid history | No typhoid history |
| **All areas** | 16.5 (15.5- 17.6) | 14.9 (13.9- 16.1) | 20.5 (15.2-27.8) | 15.9 (15.1- 16.7) |
| **Dhaka** | 33.9 (28.2- 40.8) | 32.3 (26.5-39.3) | - | - |
| **Mirzapur** | 20.5 (17.5- 23.9) | 18.5 (15.8- 21.7) | - | - |
| **Chattogram** | 16.9 (14.5- 19.6) | 17.6 (13.9- 22.2) | - | - |
| **Dinajpur** | 15.9 (13.4- 19.0) | 12.6 (10.4- 15.4) | - | - |
| **Sylhet** | 14.0 (11.6- 16.9) | 11.4 (9.5- 13.7) | - | - |
| **Satkhira** | 12.4 (10.6- 14.6) | 10.1 (8.1- 12.4) | - | - |
| **Faridpur** | 10.4 (8.5-12.7) | 11.8 (9.7- 14.3) | - | - |

\*Bangladesh-specific pooled antibody kinetics, derived from combined data for the under 5 and over 5 years age groups, were used for stratified seroincidence estimation.
\*\*95% CI, 95% confidence interval

**Supplementary Figure 1.**
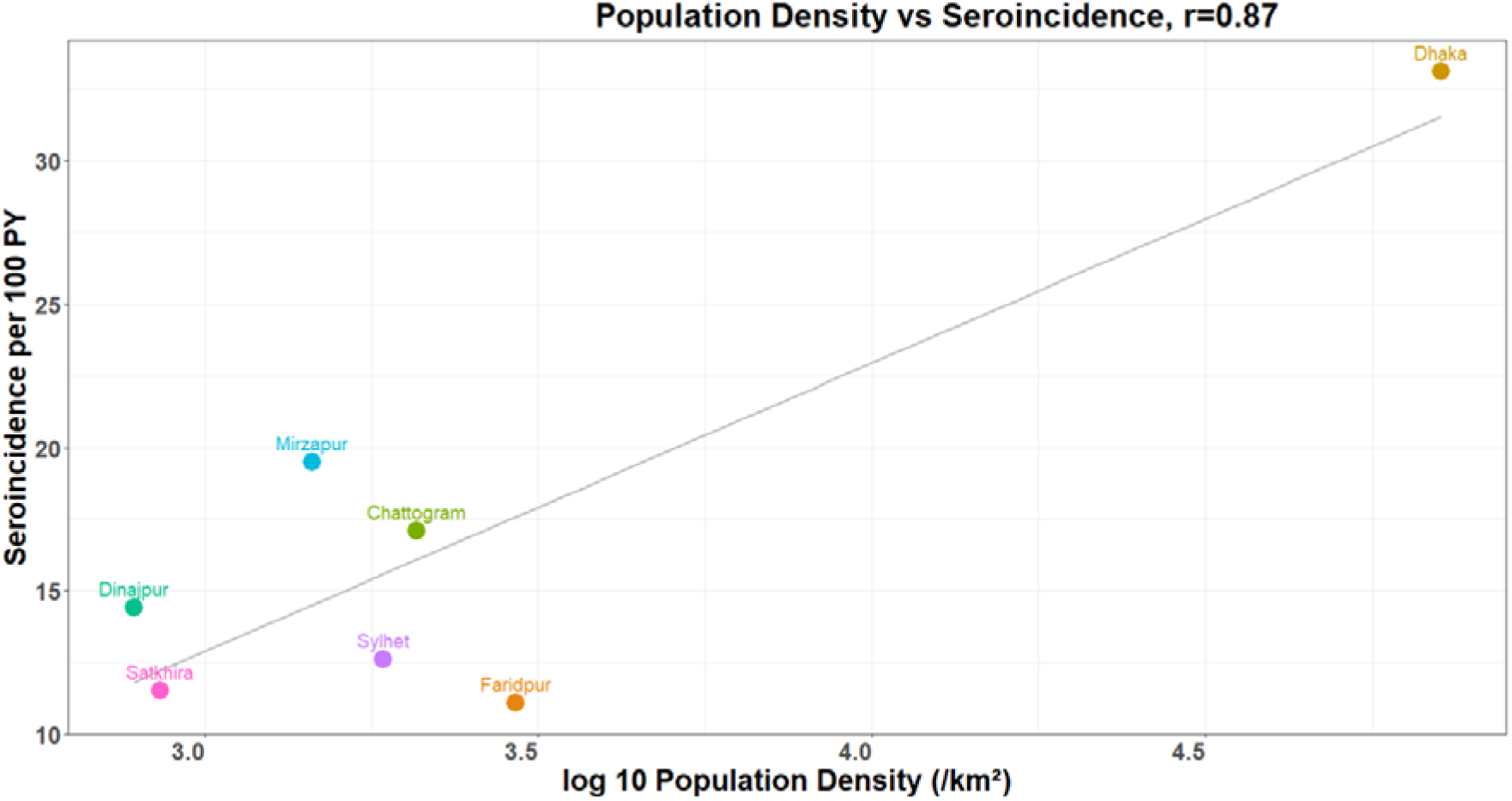
Comparison of seroincidence estimates and population densities. Each dot represents the seroincidence rate per 100 person-years plotted against log-transformed population density (people/km²) across seven areas; r denotes the Pearson correlation coefficient.

**Supplementary Figure 2.**
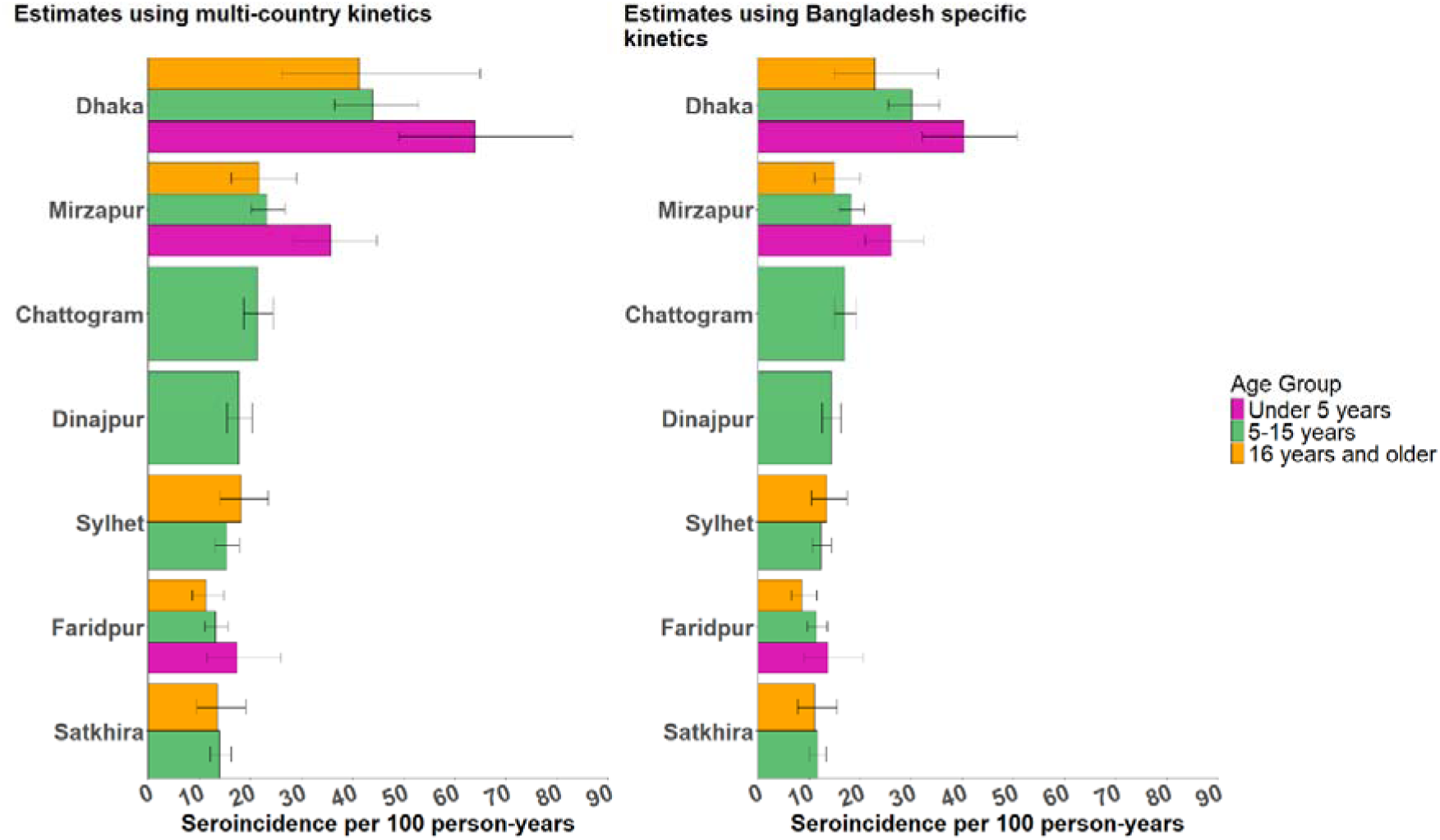
Comparison of age-stratified seroincidence estimates using multi-country versus Bangladesh-specific antibody kinetics across seven areas. Each box represents the seroincidence estimate per 100 person-years with a 95% confidence interval for a specific age group, with colours distinguishing between age groups.

## References

1. Stanaway JD, Reiner RC, Blacker BF, et al. The global burden of typhoid and paratyphoid fevers: a systematic analysis for the Global Burden of Disease Study 2017. Lancet Infect Dis 2019; 19:369–381.

2. Piovani D, Figlioli G, Nikolopoulos GK, Bonovas S. The global burden of enteric fever, 2017-2021: a systematic analysis from the global burden of disease study 2021. EClinicalMedicine 2024; 77. Available at: 10.1016/j.eclinm.2024.102883.

3. Garrett DO, Longley AT, Aiemjoy K, et al. Incidence of typhoid and paratyphoid fever in Bangladesh, Nepal, and Pakistan: results of the Surveillance for Enteric Fever in Asia Project. Lancet Glob Health 2022; 10:e978–e988. Available at: 10.1016/S2214-109X(22)00119-X.

4. Saha S, Ishtiaque Sayeed KM, Saha S, et al. Hospitalization of Pediatric Enteric Fever Cases, Dhaka, Bangladesh, 2017-2019: Incidence and Risk Factors. Clinical Infectious Diseases 2020; 71:S196–S204.

5. Naheed A, Ram PK, Brooks WA, et al. Burden of typhoid and paratyphoid fever in a densely populated urban community, Dhaka, Bangladesh. International Journal of Infectious Diseases 2010; 14:e93–e99. Available at: 10.1016/j.ijid.2009.11.023.

6. Antillon M, Saad NJ, Baker S, Pollard AJ, Pitzer VE. The Relationship between Blood Sample Volume and Diagnostic Sensitivity of Blood Culture for Typhoid and Paratyphoid Fever: A Systematic Review and Meta-Analysis. Journal of Infectious Diseases 2018; 218:S255–S267.

7. Voysey M, Pant D, Shakya M, et al. Under-detection of blood culture-positive enteric fever cases: The impact of missing data and methods for adjusting incidence estimates. PLoS Negl Trop Dis 2020; 14:1–12.

8. Bobrovitz N, Arora RK, Cao C, et al. Global seroprevalence of SARS-CoV-2 antibodies: A systematic review and meta-analysis. PLoS One 2021; 16:e0252617.

9. Arnold BF, Martin DL, Juma J, et al. Enteropathogen antibody dynamics and force of infection among children in low-resource settings. Elife 2019; 8.

10. Salje H, Cummings DAT, Rodriguez-Barraquer I, et al. Reconstruction of antibody dynamics and infection histories to evaluate dengue risk. Nature 2018; 557:719–723.

11. Kshatri JS, Bhattacharya D, Praharaj I, et al. Seroprevalence of SARS-CoV-2 in Bhubaneswar, India: findings from three rounds of community surveys. Epidemiol Infect 2021; 149:e139.

12. Watson CH, Baker S, Lau CL, et al. A cross-sectional seroepidemiological survey of typhoid fever in Fiji. PLoS Negl Trop Dis 2017; 11:1–17.

13. Meiring JE, Shakya M, Khanam F, et al. Burden of enteric fever at three urban sites in Africa and Asia: a multicentre population-based study. Lancet Glob Health 2021; 9:e1688–e1696. Available at: 10.1016/S2214-109X(21)00370-3.

14. Pulickal AS, Gautam S, Clutterbuck EA, et al. Kinetics of the natural, humoral immune response to Salmonella enterica serovar Typhi in Kathmandu, Nepal. Clinical and Vaccine Immunology 2009; 16:1413–1419.

15. Charles RC, Liang L, Khanam F, et al. Immunoproteomic analysis of antibody in lymphocyte supernatant in patients with typhoid fever in Bangladesh. Clinical and Vaccine Immunology 2014; 21:280–285.

16. Andrews JR, Khanam F, Rahman N, et al. Plasma Immunoglobulin A Responses Against 2 Salmonella Typhi Antigens Identify Patients with Typhoid Fever. Clinical Infectious Diseases 2019; 68:949–955.

17. Aiemjoy K, Seidman JC, Saha S, et al. Estimating typhoid incidence from community-based serosurveys: a multicohort study. Lancet Microbe 2022;

18. Aiemjoy K, Rumunu J, Hassen JJ, et al. Seroincidence of Enteric Fever, Juba, South Sudan. Emerg Infect Dis 2022; 28:2316–2320.

19. Abraham D, Kathiresan L, Sasikumar M, et al. Wastewater surveillance for Salmonella Typhi and its association with seroincidence of enteric fever in Vellore, India. PLoS Negl Trop Dis 2025; 19:e0012373-. Available at: 10.1371/journal.pntd.0012373.

20. Khan A, Kamenskaya P, Rezende I, et al. Seroincidence rate of typhoidal Salmonella in children in Kenya. medRxiv 2025; :2025.06.24.25330223. Available at: http://medrxiv.org/content/early/2025/06/25/2025.06.24.25330223.abstract.

21. Andrews JR, Vaidya K, Saha S, et al. Healthcare Utilization Patterns for Acute Febrile Illness in Bangladesh, Nepal, and Pakistani]: Results from the Surveillance for Enteric Fever in Asia Project. 2020; 71.

22. Saha S, Kanon N, Islam MS, et al. Cohort Profile: Health and Demographic Surveillance System in Mirzapur, Tangail, Bangladesh. medRxiv 2025; :2025.07.21.25331957. Available at: http://medrxiv.org/content/early/2025/07/22/2025.07.21.25331957.abstract.

23. Weyant C, Hooda Y, Munira SJ, et al. Cost-effectiveness and public health impact of typhoid conjugate vaccine introduction strategies in Bangladesh. Vaccine 2024; Available at: https://linkinghub.elsevier.com/retrieve/pii/S0264410X24003220. Accessed 29 March 2024.

24. Charles RC, Sultana T, Alam MM, et al. Identification of Immunogenic Salmonella enterica Serotype Typhi Antigens Expressed in Chronic Biliary Carriers of S. Typhi in Kathmandu, Nepal. PLoS Negl Trop Dis 2013; 7:1–8.

25. Teunis PFMM, van Eijkeren JCHH. Estimation of seroconversion rates for infectious diseases: Effects of age and noise. Stat Med 2020; 39:2799–2814.

26. Kristen Aiemjoy, Ezra Morrison, Kristina Lai, Jessica Siedman. Serocalculator Data Repository. 2025. Available at: https://osf.io/ne8pc/overview. Accessed 16 December 2025.

27. Lai KW, Orwa C, Seidman JC, et al. serocalculator, an R package for estimating seroincidence from cross-sectional serological data. medRxiv 2025; :2025.06.04.25328941. Available at: http://medrxiv.org/content/early/2025/06/06/2025.06.04.25328941.abstract.

28. Walker J, Russell P, Kermack L, et al. Leveraging paired serology to estimate the incidence of typhoidal *Salmonella*; infection in the STRATAA study. medRxiv 2025; :2025.03.15.25324021. Available at: http://medrxiv.org/content/early/2025/03/17/2025.03.15.25324021.abstract.

29. Hancuh M, Walldorf J, Minta AA, et al. Typhoid Fever Surveillance, Incidence Estimates, and Progress Toward Typhoid Conjugate Vaccine Introduction - Worldwide, 2018-2022. MMWR Morb Mortal Wkly Rep 2023; 72:171–176.

30. Saha S, Tanmoy AM, Andrews JR, et al. Evaluating PCR-based detection of Salmonella typhi and paratyphi a in the environment as an enteric fever surveillance tool. American Journal of Tropical Medicine and Hygiene 2019; 100:43–46.

31. Maude RR, Ghose A, Samad R, et al. A prospective study of the importance of enteric fever as a cause of non-malarial febrile illness in patients admitted to Chittagong Medical College Hospital, Bangladesh. BMC Infect Dis 2016; 16.

32. Crump JA, Luby SP, Mintz ED. The global burden of typhoid fever. 2004; 002295.

## References

1. Aiemjoy K, Seidman JC, Saha S, et al. Estimating typhoid incidence from community-based serosurveys: a multicohort study. Lancet Microbe 2022;

2. Andrews JR, Khanam F, Rahman N, et al. Plasma Immunoglobulin A Responses Against 2 Salmonella Typhi Antigens Identify Patients with Typhoid Fever. Clinical Infectious Diseases 2019; 68:949–955.

3. Charles RC, Sultana T, Alam MM, et al. Identification of Immunogenic Salmonella enterica Serotype Typhi Antigens Expressed in Chronic Biliary Carriers of S. Typhi in Kathmandu, Nepal. PLoS Negl Trop Dis 2013; 7:1–8.

4. Harris JB, LaRocque RC, Chowdhury F, et al. Susceptibility to Vibrio cholerae infection in a cohort of household contacts of patients with cholera in Bangladesh. PLoS Negl Trop Dis 2008; 2.

5. Teunis PFMM, van Eijkeren JCHH. Estimation of seroconversion rates for infectious diseases: Effects of age and noise. Stat Med 2020; 39:2799–2814.

6. Leonard MM, Camhi S, Huedo-Medina TB, Fasano A. Celiac disease genomic, environmental, microbiome, and metabolomic (CDGEMM) study design: Approach to the future of personalized prevention of celiac disease. Nutrients. 2015; 7:9325–9336.

7. James F. Pierson-Perry, Jeffrey E. Vaks, A. Paul Durham, et al. EP17 _ Evaluation of Detection Capability for Clinical Laboratory Measurement Procedures. 2012: 80. Available at: https://clsi.org/standards/products/method-evaluation/documents/ep17/. Accessed 23 July 2025.

8. Weyant C, Hooda Y, Munira SJ, et al. Cost-effectiveness and public health impact of typhoid conjugate vaccine introduction strategies in Bangladesh. Vaccine 2024; Available at: https://linkinghub.elsevier.com/retrieve/pii/S0264410X24003220. Accessed 29 March 2024.

